# Two pathways to self-harm in adolescence

**DOI:** 10.1101/2020.07.10.20150789

**Authors:** Stepheni Uh, Edwin S. Dalmaijer, Roma Siugzdaite, Tamsin J. Ford, Duncan E. Astle

**Author notes:** **Corresponding author:** Stepheni Uh, MRC Cognition and Brain Sciences Unit, University of Cambridge; 15 Chaucer Road, CB2 7EF, United Kingdom.

## Abstract

**Background:** The behavioural and emotional profiles underlying adolescent self-harm, and its developmental risk factors, are relatively unknown. We aimed to identify sub-groups of young people who self-harm (YPSH) and longitudinal predictors leading to self-harm.

**Methods:** Participants were from the Millennium Cohort Study (n=10,827). A clustering algorithm identified sub-groups who self-harmed with different behavioural and emotional profiles at age 14. Feature selection analyses were then used to identify longitudinal predictors of self-harming behaviour.

**Findings:** There were two distinct sub-groups at age 14: a smaller group (n = 379) who reported a long history of psychopathology, and a second group (n = 905) without. Notably, both groups could be predicted almost a decade before the reported self-harm. They were similarly characterised by sleep problems and low self-esteem, but there was developmental differentiation. From an early age, the first group had poorer emotion regulation, were bullied, and their caregivers faced emotional challenges. The second group showed less consistency in early childhood, but later reported more willingness to take risks and less security with peers/family.

**Interpretation:** Our results uncover two distinct pathways to self-harm: a ‘psychopathology’ pathway, associated with early and persistent emotional difficulties and bullying; and an ‘adolescent risky behaviour’ pathway, where risk-taking and external challenges emerge later into adolescence and predict self-harm. These two pathways have long developmental histories, providing an extended window for interventions as well as potential improvements in the identification of children at risk, biopsychosocial causes, and treatment or prevention of self-harm.

**Funding:** This study was supported by the UK Medical Research Council, Templeton World Charitable Foundation, and a Gates Cambridge Scholarship awarded to SU.

## Introduction

Self-harm is commonly associated with poor mental health in both clinical and non-clinical populations, with prevalence estimates ranging from approximately 13·2 to 19·7% among adolescents in England.^1–3^ The definition of self-harm varies, due to complexity in its presentation and description (e.g., nonsuicidal self-harm/self-injury,^4^ deliberate self-harm^5^). For the present purposes it is defined as the purposeful act of hurting oneself with or without suicidal intent. Self-harm is a significant risk factor for subsequent suicide attempts, and consequently, is a strong predictor of death by suicide among adolescents^6^ and other harmful outcomes including risky behaviours like substance abuse.^1,6^

Despite its prevalence and lifelong consequences, there has been little progress in the accurate prediction of self-harm,^7^ partly due to the unclear interaction(s) between the individual and external risk factors that predict it.^8^ A study on adolescents in the UK found that repeated self-harm was strongly linked to personality disturbances, depression, substance use, troubled relationships with peers/family, poor school performance, and chronic psychosocial as well as behavioural problems.^9^ Studies also indicate that socioeconomic factors predict self-harm and suicidal behaviours in adults and adolescents.^10,11^ In short, there are likely internal and external risk factors for self-harm.

An added complication is that there may be different *subtypes* of self-harming behaviour, with distinct sets of risk factors. One study suggests that there are two subtypes of self-harm: (1) occasional self-harm linked to social factors like academic achievement and family related problems while (2) repetitive self-harm was more associated with internal factors such as prior suicidal ideation, body-image issues, as well as anxiety and depressive symptoms.^12^ In addition to different types of self-harm, there is also the possibility of different sub-groups of *young people who self-harm* (YPSH). For instance, recent research reports that there may be distinct psychological profiles of YPSH based on self-report questionnaires covering traits like self-esteem, depression, and impulsivity.^13,14^

Despite the increase in self-harm research over recent decades, we have made minimal progress in addressing a key set of questions, relevant to researchers, policy makers, and practitioners.^15^ What are the risk factors for self-harm? Will all YPSH present with a similar emotional and behavioural profile? And finally, how early in childhood do these risks emerge? The purpose of this study is to address these questions.

Previous work exploring self-harm has largely focused on those who present in hospital or other clinical facilities,^1,3^ but this is unlikely to capture all self-harming behaviour. McManus and colleagues,^1^ for instance, found that self-reported lifetime non-suicidal self-harm increased from 2.4% in 2000 to 6.4% in 2014 in England, but most did not present to medical or psychological services. This highlights that those who seek help after harming themselves are likely to vary in a number of ways from those who do not seek help; thus, predictors identified in clinically recruited samples may not be generalizable to YPSH in the population. Another limitation of relying solely on clinical recruitment is that we are unable to investigate the early developmental trajectories of adolescents who ultimately self-harm in comparison to those who do not.^16^

In the current study, we identified adolescents who reported self-harm at age 14, from a nationally representative UK birth cohort of approximately 19,000 individuals. We then used a machine learning analysis to identify whether there are distinct clusters of YPSH, with different emotional and behavioural presentations. We subsequently identified the concurrent risk factors using the extensive dataset available on these individuals. Finally, we used the preceding waves of data collection from when the children were 5, 7 and 11 years of age to identify risk factors from early and middle childhood.

## Methods

### Participants

The Millennium Cohort Study^17^ (MCS) is a large-scale ongoing longitudinal developmental study of young people throughout the UK. An extensive amount of behavioural, socio-emotional, and physical data on the participants has been collected since they were 9 months old. From the original derived dataset of 11,884 individuals at age 14, we included 10,827 (50% female) participants who had complete responses to the measures used in our analysis. Within the participants who reported self-harm (n = 1580, 73% female), our main analysis would subsequently focus on a large subset (n = 1284, 74% female) who were identified as members of two distinct behavioural clusters. We also included a random sub-sample of participants (n = 900, 70% female) who did not self-harm, as a comparison group for subsequent analyses. For validation purposes, a “train” sample (70% of total) was taken from each group and used to fit our model while each held-out “test” sample (30%) was used to validate the predictors. There was slight variation in numbers in each sweep due to small differences in missing predictor data, which will be reported alongside the results.

### Measures

The Strengths and Difficulties Questionnaire (SDQ) is a validated 25-item screening measure of children’s (ages 3–16) mental health and behavioural problems utilized in both clinical and research settings.^18^ In this study, caregiver-completed SDQ data was collected at ages 5, 7, 11, and 14 – sweeps three, four, five, and six, respectively. The Mood and Feelings Questionnaire (MFQ), a reliable and validated self-report measure of depressive feelings and behaviours in children and adolescents (ages 6–17),^19^ was completed at age 14. Finally, the item “In the past year have you hurt yourself on purpose?”, administered at age 14, was used as an index of self-harm. The SDQ, MFQ, and self-harm item were used in data-driven analyses to describe the emotional profile(s) of self-harmers.

A large number (78-100 per sweep) of potential predictor variables was selected from the MCS dataset on the basis of the previous literature (see appendix). We grouped these variables according to six domains that have been broadly discussed across prior self-harm research: child health (e.g., sleep, alcohol consumption); child mental health (e.g., emotion issues, self-esteem); caregiver mental health (e.g., health limitations); home environment (e.g., housing tenure, neighbourhood safety); peer relations (e.g., quality of friendships); and adversity (e.g., bullying).

### Statistical analysis

Our first goal was to characterise the profiles of YPSH at age 14. To do this we entered z-scored SDQ and MFQ data to a simple artificial neural network.^20^ This is an unsupervised machine learning algorithm that learned about different profiles of scores across measures in the *N* = 10,827 dataset. This type of non-linear data reduction technique is ideally suited to high- dimensional datasets because, unlike other data reduction techniques, it does not group variables or identify latent factors. Instead, it preserves information about potentially distinct profiles within the dataset, captures non-linear relationships, and allows for measures to be differentially related across the sample.^21–23^ We then employed k-means clustering to determine whether different sub-groups of YPSH existed and how members of those sub-groups differed from each other.

In order to identify longitudinal and concurrent predictors for sub-group membership, we utilised logistic regression with regularisation through Least Absolute Shrinkage and Selection Operator (LASSO)^24^ on our train samples. LASSO is a supervised algorithm typically used for feature selection regularisation. It models the outcome (i.e., self-harm sub-group versus comparison), while emphasising the most important predictors by shrinking coefficients of less important predictors to zero. In each group, participants missing more than 30% of their data were excluded, while those with less than 30% missing data had missing values imputed through k-nearest-neighbour imputation (k = 25).

Within the train sample, each sweep was subjected to 1000 bootstrapping iterations of logistic regression with LASSO regularisation and 5-fold cross-validation. As this procedure selects stronger predictors while down-weighting weaker predictors within each iteration, we selected only those predictors that were non-zero in 95% of iterations. Finally, to verify their predictive accuracy, the features selected using this cross-validation within the training sample were used to predict self-harm in the test sample through standard logistic regression. Only predictors that survived all of these steps, including the final validation on the test samples, were considered genuine predictors of self-harm behaviour.

### Role of the funding source

The funder had no role in the study design, data collection, data analysis, data interpretation, or writing of the report. The corresponding author had full access to all the data in the study and had final responsibility for the decision to submit for publication.

## Results

Our artificial neural network and clustering analysis on the resulting node weights identified two clusters of YPSH: one with high SDQ and MFQ scores reflecting psychopathology (n = 379, labelled “Group 1”) and one with age-appropriate scores reflecting lack of psychopathology (n = 905, labelled “Group 2”). The silhouette coefficient, a validity measure for cluster analyses, was 0.52 for this solution. This indicates good separation between the two communities of YPSH.^25^ Table 1 shows descriptive information for all group samples.

**Table 1:** Descriptive information for the complete case samples.

|  |  | **Group 1**<br>**(n=379)** | **Group 2**<br>**(n = 905)** | **Comparison**<br>**(n = 900)** |
| --- | --- | --- | --- | --- |
| **Sex** |  |  |  |  |
|  | Female | 269 (71%) | 686 (76%) | 630 (70%) |
|  | Male | 110 (29%) | 219 (24%) | 270 (30%) |
| **Ethnicity** |  |  |  |  |
|  | White | 349 (92%) | 822 (91%) | 790 (88%) |
|  | Mixed | 11 (3%) | 29 (3%) | 18 (2%) |
|  | Indian | 3 (1%) | 13 (1%) | 19 (2%) |
|  | Pakistani and Bangladeshi | 8 (2%) | 21 (3%) | 44 (5%) |
|  | Black or Black British | 5 (1%) | 12 (1%) | 22 (2%) |
|  | Other Ethnic group | 3 (1%) | 8 (1%) | 7 (1%) |
| **OECD below 60% median**<br>**poverty** |  |  |  |  |
|  | Age 5 | 179 (47%) | 217 (24%) | 206 (23%) |
|  | Age 7 | 157 (41%) | 179 (20%) | 197 (22%) |
|  | Age 11 | 127 (34%) | 125 (14%) | 145 (16%) |
|  | Age 14 | 148 (39%) | 186 (21%) | 177 (20%) |

Data are n (%). Numbers vary because of missing data

We initially tracked the SDQ scores back through earlier sweeps, to test whether these differences are consistent across developmental time (Figure 1). A two-factor mixed ANOVA, Greenhouse-Geisser corrected, showed a significant interaction between group membership (Group 1 (with psychopathology), Group 2 (without psychopathology), and Comparison) and age (Ages 5, 7, 11, 14) for all SDQ subdomain scores (Emotional symptoms: F(5·52, 6020·70) = 96·20, p < 0·0001; Conduct problems: F(5·64, 6149·33) = 47·23, p < 0·0001; Hyperactivity/Inattention: F(6·65, 6059·84 = 27·92), p < 0·0001; Peer relationships problems: F(5·55, 6054·50) = 66·44, p < 0·0001; Prosocial behaviours: F(5·48, 5980·45) = 33·67, p < 0·0001). These interactions result from the increasing psychopathology for Group 1 and their increasing divergence from the other two groups growing over developmental time (versus Group 2: all Fs > 51·64, all ps < 0·0001; versus Comparison sample: all Fs > 36·78, all ps < 0·05), whereas the trajectories for the other two groups do not differ except for emotional symptoms (Group 2 versus Comparison: Emotional symptoms (F(2·77, 4989·98) = 3·680), p = 0·014; all other Fs > 0·31, all ps > 0·05).

**Figure 1:**
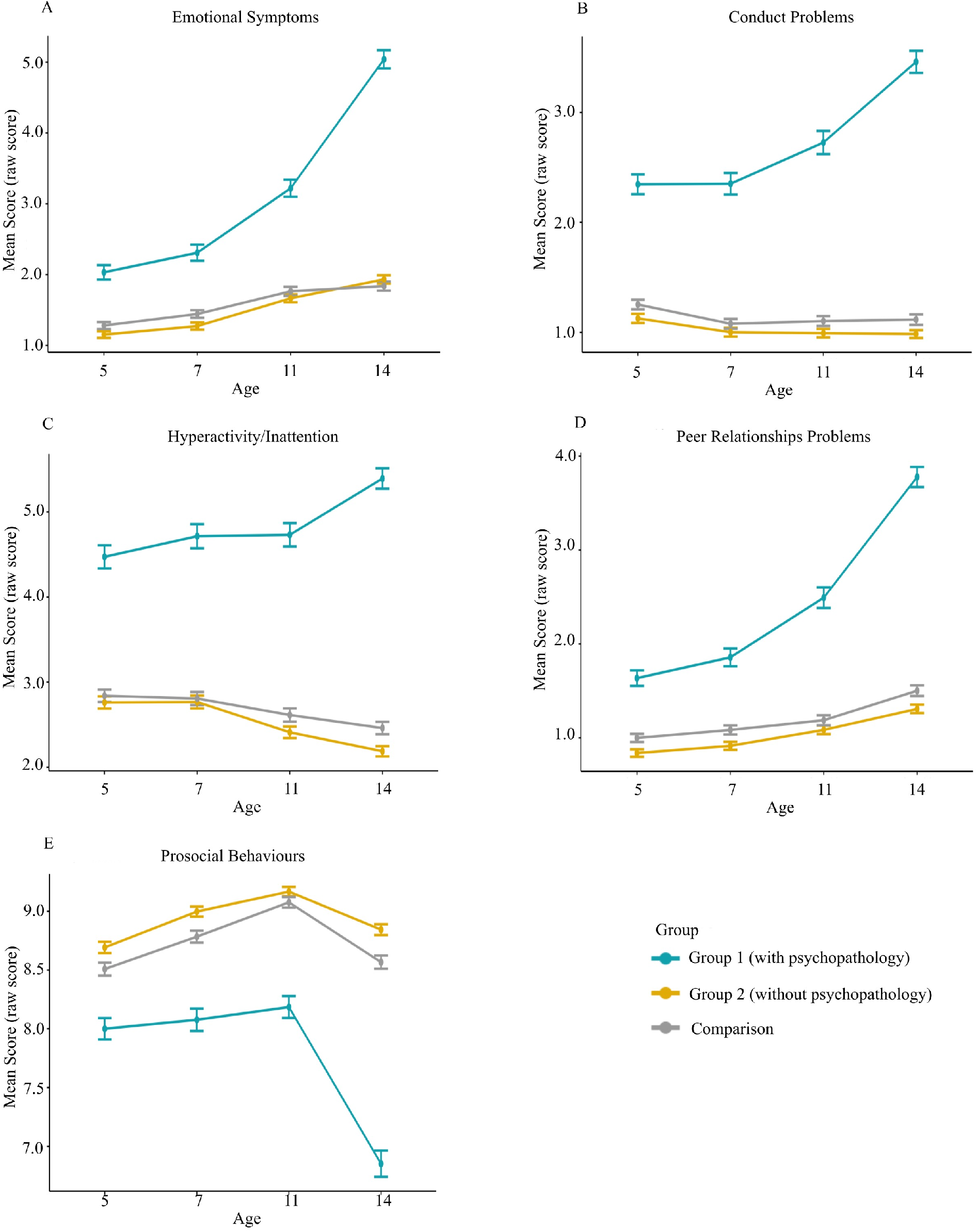
The developmental trajectories from ages 5-14 for Group 1, Group 2, and the comparison group of mean SDQ subdomain raw scores: A) Emotional symptoms, B) Conduct problems, C) Hyperactivity/inattention, D) Peer relationships problems, and E) Prosocial behaviours. Error bars show the 95% CIs.

After applying the cross-validated LASSO on the train set and then testing the surviving predictors in the test set, we identified concurrent self-harm predictors at age 14, when participants answered the self-harm index (Figure 2D). The strengths of the validated predictors for both groups were assessed by the absolute values of the standardized coefficients computed from the LASSO procedure. The strongest predictors for Group 1 (with psychopathology; n = 249, 73% female) were poor emotional control (β = 0·70), low self-esteem (β = 0·50), waking during sleep (β = 0·22), trouble falling asleep (β = 0·21), more quarrels with caregivers (β = 0·14), and being unhappy at school (β = 0·14). Predictors for Group 2 (without psychopathology; n = 614, 77% female) were low self-esteem (β = 0·57), low support system from peers/family (β = 0·16), trouble falling asleep (β = 0·09), being more willing to take risks (β = 0·08), and having caregivers with self-reported higher extraversion (β = 0·07).

**Figure 2:**
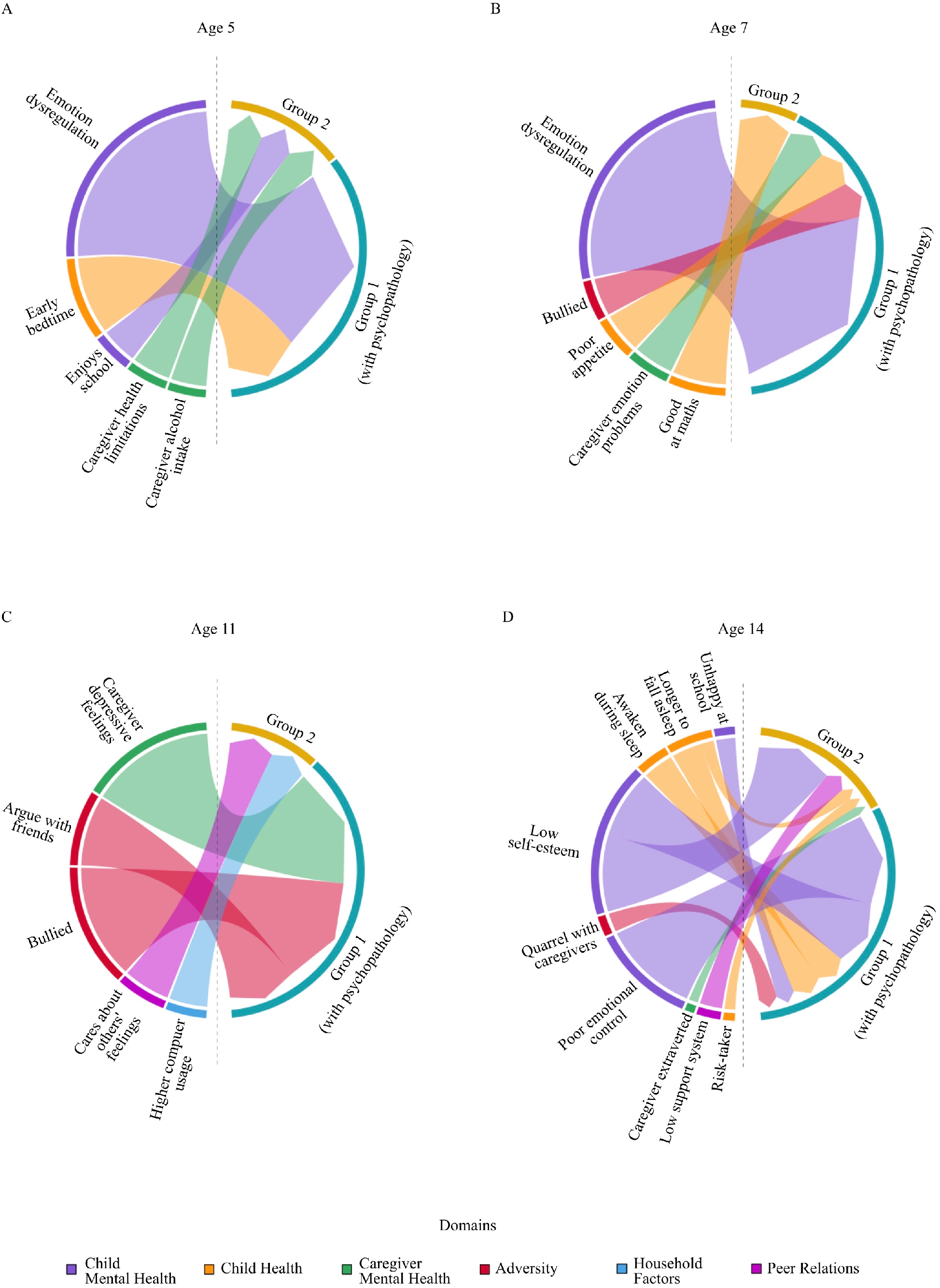
Predictors across six domains of self-harm for Group 1 (with psychopathology) and Group 2 membership (without psychopathology) reported A) from age 5, B) from age 7, C) from age 11, and D) at age 14.

Longitudinal predictors were identified at ages 5, 7, and 11 (Figure 2A–C). At age 11, later membership of Group 1 (with psychopathology; n = 251, 73% female) was predicted by caregiver depressive feelings (β = 0·26, p = 0·052), being bullied (β = 0·24), and frequently arguing with friends (β = 0·15). Later membership of Group 2 (n = 608, 76% female) was predicted by caring about the feelings of others (β = 0·10) and more weekday hours spent on the computer/games (β = 0·08). At age 7, the predictors for Group 1 (n = 271, 73% female) were child emotion dysregulation issues (β = 0·40), poorer appetite (β = 0·07), caregiver emotional problems (β = 0·07), and being bullied (β = 0·07). Group 2 membership (n = 644, 76% female) was predicted by having less difficulty with maths (β = 0·09) at age 7. Lastly, at age 5, predictors of later Group 1 membership (n = 271, 73% female) were child emotion dysregulation issues (β = 0·28) and earlier weekday bedtimes (β = 0·10), while later Group 2 membership (n = 644, 76% female) was predicted by caregiver health limitations on work (β = 0·05), child enjoying school (β = 0·05), and more frequent caregiver alcohol intake (β = 0·05).

In summary: we identified distinct sub-groups among YPSH, with significant risk factors present as early as age 5, nearly a decade before they report self-harming. While sleep difficulties and low self-esteem reported at age 14 commonly predicted self-harm behaviour, irrespective of sub- group, there were divergences in other predictors.

## Discussion

### Two sub-groups among young people who self-harm

This study used a data-driven approach to identify two distinct sub-groups among YPSH in a nationally representative cohort. Crucially, the membership of these groups could be predicted 9 years earlier. The smaller sub-group (Group 1) presented with psychopathological traits, in comparison to the larger sub-group (Group 2) who had age-appropriate scores on both the SDQ and MFQ. The longitudinal nature of our analysis allowed us to distinguish characteristics that appear *alongside* self-harm behaviour (e.g., low self-esteem) from those that *precede* it (e.g., bullying). The pathway for Group 1 self-harmers embodies a ‘psychopathology’ route, in which a long history of emotional dysregulation, psychopathology, and bullying precedes self-harm. Group 2, meanwhile, do not fit this profile, or that suggested by previous research.^14^ Their self-harm behaviour is harder to predict early in childhood and instead coincides with later increases in risk-taking and changes in their relationships with family and friends – the ‘adolescent risky behaviour’ pathway. The two groups we found are in line with recent research findings of distinct profiles of YPSH,^13,14^ which is encouraging. Interestingly, Stanford and colleagues^13,14^ found a separate “impulsive” profile of self-harmers, while we found that risk-taking was a concurrent predictor for the larger non-pathological group.

### Shared and distinct risk factors for adolescent self-harm

Both caregiver and self-report measures were used to determine the emotional, behavioural, and mental health profiles of YPSH and their consistency across development. Those with a psychopathological profile in Group 1 were reported by their caregivers to have emotional and behavioural difficulties as early as age 5, which gradually deteriorated over time. They also self- reported poor mental health at age 14 when they reported self-harm. Caregivers of young people in Group 2, meanwhile, reported little to no psychological distress in childhood or adolescence. Furthermore, the participants themselves indicate good mental health – so it is not simply the case that caregivers are unaware of underlying mental health difficulties. In essence, Group 2 does not present, or even self-report, the psychopathological traits that have been most associated with those who self-harm, unlike their Group 1 counterparts. This group fits the non-psychopathological profile reported by Stanford and colleagues.^14^ However, they are indeed distinct from the comparison sample, as our subsequent analysis of concurrent and preceding risk factors would reveal.

Low self-esteem and sleep problems – significant risk factors for suicidal behaviours and poorer mental health^16^ – were shared concurrent predictors for both sub-groups. However, their respective concurrent and longitudinal predictors were mostly distinct. Young people in Group 1 (with psychopathology) were more strongly associated with mental health challenges in addition to adverse relationships with caregivers at age 14. Longitudinally, greater emotion dysregulation, being bullied, and caregiver mental health issues (e.g., emotional problems) were significant predictors throughout their development: common risk factors for self-harm.^26^ Importantly, the longitudinal perspective design highlights that this psychopathology route to self-harm starts early, with its origins in poor mental health for both children and their caregivers.

Concurrent predictors for the second, larger, non-psychopathological group of YPSH were feeling insecure with their peers and family (low support), being more willing to take risks, and having more extraverted caregivers. Risk-taking, in particular, has been empirically and conceptually linked to self-harm as both are subject to peer influence^27^ and impulsivity^28,29^: factors that may limit time spent considering alternate coping methods and the consequences of self-harm.^14^ Longitudinal predictors, meanwhile, were less associated with mental health challenges and less consistent from an early age. At age 11, for instance, the strongest predictor suggested that they were more concerned about the feelings of others alongside greater usage of computers and games. Interestingly, at ages 5 and 7, caregivers reported positive school-related predictors, but neither was particularly strong. Hypothetically, these YPSH do not externalise their difficulties, especially as they do not seem to feel safe with their family/friends. They may also find it difficult to connect with their caregivers’ extraversion, who therefore are unaware of their struggles – which may explain the case of why many self-harm incidents are unknown by caregivers.^30^ Overall, the predictors for the ‘adolescent risky behaviour pathway’ appear to be more representative of later external experiences along with the emergence of their willingness to engage in risky behaviours and feeling less security with peers and family.

### From theory to practice: implications for policymakers and practitioners

A key implication of our findings is that we have a decade-long window to intervene for some children who are at increased risk of self-harm as adolescents. Early targeted interventions, particularly those focused on emotion regulation, may be helpful for this group. A meta-analysis on resilience interventions in schools highlights that effectiveness can depend on age and mental health outcomes; for instance, childhood interventions are relatively effective for general psychological distress.^31^ The persistence of psychopathology among Group 1, furthermore, suggests that early screening measures if combined with prompt access to effective intervention could reduce the risk of future self-harm as well as improve mental health in the short term.^7^ A second and highly tractable target for intervention is bullying, which casts a shadow over adult as well as childhood mental health.^32^ This was a strong and early predictor of self-harm for children in the psychopathology pathway, preceding self-harm reports by 7 years. There are now a number of evidence-based anti-bullying interventions that can be deployed at a school level that could and should be implemented.^33,34^

The larger Group 2, without psychopathology, represents the challenge we face to assist those in the general population.^14^ Access to universal programmes and materials for self-help and problem-solving/conflict regulations (as recommended for inclusion in PSHE education^35^) may be effective for those who do not seek help from formal services. Sleep training is also an area to consider. Sleep difficulties were strong overlapping concurrent predictors for self-harming behaviour in our study and has been associated with emotion regulation and mood disorders^36^ as well as increased suicide risk.^16^ Additionally, targeted interventions by mental health leaders and school-based mental health teams are important. Training for teachers, especially, could be critical as they are often the first people to hear about self-harm but may have difficulty responding.

### Future Directions

As there is a lack of research investigating early childhood origins of self-harm^7,37^ and sub- groups among YPSH, our study provides an important foundation for future research. An essential future direction is to replicate our findings in other national and international cohorts, particularly as our work only partially replicated that of previous groups.^13,14^ Thus, a next step is to further refine psychological sub-groups of YPSH. Where anti-bullying programmes are trialled or implemented, permission to link to administrative data (e.g., health records) would allow future research to explore presentations for self-harm among the cohort in adolescents. Targeted prevention for substance misuse has successfully applied personality measures to tailor school-based programmes to pupils’ needs; our findings suggest that a similar approach may be worth considering among pre-teens in relation to self-harm.^38^

### Study limitations

The longitudinal analysis of a nationally representative sample along with powerful predictive analyses, using machine learning, provides valuable insight into the developmental pathways leading to self-harm. Nevertheless, several limitations to this study exist. First, the self-harm index used in this study is a binary yes-or-no response despite the complex nature and range in severity of self-harm.^5^ The way in which participants hurt themselves or more probing questions into the motivations behind this behaviour were not collected. Despite not capturing these nuances, the simple measure of asking whether children have self-harmed in the previous year still provided a concrete window into a mental health problem that is extremely difficult to measure accurately and in detail, particularly given the large sample size. Second, we intentionally did not include the sex of the participants as a predictor. Instead we incorporated the fact that approximately 70-77% of the self-harmers were female in our matched comparison sample. This pattern is well-established^37^ and even in a population sample, separate analyses by sex would suffer from low statistical power from which to explore self-harm among boys. Thirdly, our statistical approach was incredibly conservative (cross-validated regularisation, bootstrapping, and a final validation in a test sample) when analysing the predictors for this outcome, which may serve as both a strength and limitation of this study. It is likely that we overlooked meaningful weaker predictors, but this comes at the benefit of avoiding over-fitting and suggests that our reported findings are robust.

### Conclusions

There is global consensus that self-harm is a prevalent concern in adolescence and a priority for public health efforts. Establishing early risk factors and behavioural profiles that can be traced and tracked across development provides a crucial step towards the early identification of these young people, to elucidating underlying casual mechanisms, and ultimately prevention and treatment. We show that there are two relatively distinct profiles among adolescents who self-harm – early and persistent psychopathology and exposure to bullying versus adolescent risk taking – and that these profiles have different developmental pathways.

## Data Availability

The data is made available by the UK Data Service

## Contributors

SU, ESD, RS, and DEA contributed to the conception and design of the study. SU, ESD, and DEA contributed to the procedures and analyses of the study. SU and ESD did the analyses, and SU drafted the Article. All authors contributed to the interpretation of data and critically reviewed the writing. TJF critiqued the output for important policy and clinical implications. All authors have read and approved the final version of the manuscript.

## Declaration of interests

We declare no competing interests

## Acknowledgements

SU was supported by the Gates Cambridge Trust. ESD and DEA are supported by grant TWCF0159 from the Templeton World Charity Foundation to DEA. SU, RS, and DEA are supported by the UK Medical Research Council, grant MC-A0606-5PQ41. We are grateful to the Centre for Longitudinal Studies (CLS), UCL Institute of Education, for the use of these data and to the UK Data Service for making them available. However, neither CLS nor the UK Data Service bear any responsibility for the analysis or interpretation of these data. We also thank Emma Jayne Kilford for providing feedback on an earlier version of this manuscript.

## Notes

### Competing Interest Statement

The authors have declared no competing interest.

### Author Declarations

We received permission and access to the open source data of the Millennium Cohort Study, provided by the UK Data Service and the Centre for Longitudinal Studies (CLS), UCL Institute of Education.

## References

1 McManus S, Gunnell D, Cooper C, et al. Prevalence of non-suicidal self-harm and service contact in England, 2000–14: repeated cross-sectional surveys of the general population. The Lancet Psychiatry 2019; 0: 1–9. DOI:10.1016/S2215-0366(19)30188-9

2 Muehlenkamp JJ, Claes L, Havertape L, Plener PL. International prevalence of adolescent non-suicidal self-injury and deliberate self-harm. Child Adolesc Psychiatry Ment Health 2012; 6: 10. DOI:10.1186/1753-2000-6-10

3 O’connor RC, Rasmussen S, Miles J, Hawton K. Self-harm in adolescents: self-report survey in schools in Scotland. BJPsych 2009; 194: 68–72. DOI:10.1192/bjp.bp.107.047704

4 International Society for the Study of Self-Injury. What is self-injury? https://itriples.org/category/about-self-injury/#what-is-self-injury (accessed 30 January, 2020).

5 Chapman AL, Gratz KL, Brown MZ. Solving the puzzle of deliberate self-harm: The experiential avoidance model. Behav Res Ther 2006; 44(3): 371–394. DOI:10.1016/J.BRAT.2005.03.005

6 Castellví P, Lucas-Romero E, Miranda-Mendizábal A, et al. Longitudinal association between self-injurious thoughts and behaviors and suicidal behavior in adolescents and young adults: A systematic review with meta-analysis. J Affect Disord 2017; 215: 37–48. DOI:10.1016/j.jad.2017.03.035

7 Westers NJ, Plener PL. Managing risk and self-harm: Keeping young people safe. Clin Child Psychol Psychiatry 2019; 00(0): 1–15. DOI:10.1177/1359104519895064

8 Gratz KL. Risk factors for and functions of deliberate self-harm: An empirical and conceptual review. Clin Psychol Sci Pract 2003; 10(2): 192–205. DOI:10.1093/clipsy/bpg022

9 Hawton K, Rodham K, Evans E, Weatherall R. Deliberate self harm in adolescents: self report survey in schools in England. BMJ 2002; 325(7374): 1207–1211. DOI:10.1136/bmj.325.7374.1207

10 Ayton A, Rasool H, Cottrell D. Deliberate self-harm in children and adolescents: Association with social deprivation. Eur Child Adolesc Psychiatry 2003; 12: 303–307. DOI:10.1007/s00787-003-0344-0

11 Greydanus DE, Shek D. Deliberate self-harm and suicide in adolescents. Keio J Med 2009; 58(3): 144–151.

12 Brunner R, Parzer P, Haffner J, et al. Prevalence and psychological correlates of occasional and repetitive deliberate self-harm in adolescents. Arch Pediatr Adolesc Med 2007; 161(7): 641–649.

13 Stanford S, Jones MP. Psychological subtyping finds pathological, impulsive, and “normal” groups among adolescents who self-harm. J Child Psychol Psychiatry 2009; 50(7): 807–815. DOI:10.1111/j.1469-7610.2009.02067.x

14 Stanford S, Jones MP, Hudson JL. Rethinking pathology in adolescent self-harm: Towards a more complex understanding of risk factors. J Adolesc 2017; 54: 32–41. DOI:10.1016/j.adolescence.2016.11.004

15 Franklin JC, Ribeiro JD, Fox KR, et al. Risk factors for suicidal thoughts and behaviors: a meta-analysis of 50 years of research. Psychol Bull 2016; 134(2): 187–232. DOI:10.1037/bul0000084

16 Mars B, Heron J, Klonsky ED, et al. Predictors of future suicide attempt among adolescents with suicidal thoughts or non-suicidal self-harm: a population-based birth cohort study. The Lancet Psychiatry 2019; 6(4): 327–337. DOI:10.1016/S2215-0366(19)30030-6

17 Joshi H, Fitzsimons E. The UK Millennium Cohort Study: the making of a multi-purpose resource for social science and policy in the UK. Longit Life Course Stud 2016; 7(4): 431– 440. DOI:10.14301/llcs.v7i4.416

18 Becker A, Woerner W, Hasselhorn M, Banaschewski T, Rothenberger A. Validation of the parent and teacher SDQ in a clinical sample. Eur Child Adolesc Psychiatry [Suppl 2] 2004; 13: 11–16. DOI:10.1007/s00787-004-2003-5

19 Daviss WB, Birmaher B, Melhem NA, Axelson DA, Michaels SM, Brent DA. Criterion validity of the Mood and Feelings Questionnaire for depressive episodes in clinic and non-clinic subjects. J Child Psychol Psychiatry 2006; 47(9): 927–934. DOI:10.1111/j.1469-7610.2006.01646.x

20 Kohonen T. The Self-Organizing Map. Proc IEEE 1990; 78(9): 1464–1480.

21 Astle DE, Bathelt J, CALM team, Holmes J. Remapping the cognitive and neural profiles of children who struggle at school. Dev Sci 2019; 22(1): e12747.

22 Dittenbach M, Rauber A, Merkl D. Uncovering hierarchical structure in data using the growing hierarchical self-organizing map. Neurocomputing 2002; 48(2002): 199–216.

23 Rennie JP, Zhang M, Hawkins E, Bathelt J, Astle DE. Mapping differential responses to cognitive training using machine learning. Dev Sci 2019; e0012868.

24 Tibshirani R. Regression Shrinkage and Selection via the Lasso. J. R. Statist. Soc. B 1996; 58: 267–288.

25 Kaufman L, Rousseeuw PJ. Finding groups in data: an introduction to cluster analysis. John, John Wiley and Sons 1990.

26 Skegg K. Self-harm. Lancet 2005; 366(9495): 1471–1483. DOI:10.1016/S0140-6736(05)67600-3

27 Blakemore S-J, Mills KL. Is adolescence a sensitive period for sociocultural processing? Annu Rev Psychol 2014; 65: 187–207. DOI:10.1146/annurev-psych-010213-115202

28 Lockwood J, Daley D, Townsend E, Kapil Sayal. Impulsivity and self-harm in adolescence: a systematic review. Eur Child Adolesc Psychiatry 2017; 26: 387–402. DOI:10.1007/s00787-016-0915-5

29 Vrouva I, Fonagy P, Fearon PRM, Roussow T. The risk-taking and self-harm inventory for adolescents: Development and psychometric evaluation. Psychol Assess 2010; 22(4): 852– 865. DOI:10.1037/a0020583

30 DeVille DC, Whalen D, Breslin FJ, et al. Prevalence and family-related factors associated with suicidal ideation, suicide attempts, and self-injury in children aged 9 to 10 years. JAMA Netw open 2020; 3(2): e1920956. DOI:10.1001/jamanetworkopen.2019.20956

31 Dray J, Bowman J, Campbell E, et al. Systematic review of universal resilience-focused interventions targeting child and adolescent mental health in the school setting. J Am Acad Child Adolesc Psychiatry 2017; 56(10): 813–824. DOI:10.1016/j.jaac.2017.07.780

32 Arseneault L. Annual research review: The persistent and pervasive impact of being bullied in childhood and adolescence: implications for policy and practice. J Child Psychol Psychiatry 2018; 59(4): 405–421. DOI:10.1111/jcpp.12841

33 Department of Health & Social Care, Department for Education. Transforming children and young people’s mental health provision: a green paper. London, UK: Crown, 2017.

34 Anti-Bullying Alliance. All together: whole school anti-bullying programme. https://www.anti-bullyingalliance.org.uk/aba-our-work/our-programmes/all-together-whole-school-anti-bullying-programme (accessed 13 May, 2020).

35 School Wellbeing. PSHE: personal social health education. https://www.schoolwellbeing.co.uk/pages/pshe (accessed 13 May, 2020).

36 Van Der Helm E, Walker MP. Overnight Therapy? The Role of Sleep in Emotional Brain Processing. Psychol Bull 2009; 135(5): 731–748. DOI:10.1037/a0016570

37 Hawton K, Saunders KEA, O’Connor RC. Self-harm and suicide in adolescents. Lancet 2012; 379: 2373–2382. DOI:10.1016/S0140-6736(12)60322-5

38 Conrod PJ. Personality-targeted interventions for substance use and misuse substance misuse in north america. Curr Addict Reports 2016; 3: 426–436. DOI:10.1007/s40429-016-0127-6

